# Structural covariance networks in the fetal brain reveal altered neurodevelopment for specific subtypes of congenital heart disease

**DOI:** 10.1101/2024.01.30.24302035

**Authors:** Siân Wilson, Daniel Cromb, Alexandra F. Bonthrone, Alena Uus, Anthony Price, Alexia Egloff, Milou P.M Van Poppe, Johannes K Steinweg, Kuberan Pushparajah, John Simpson, David FA Lloyd, Reza Razavi, Jonathan O’Muircheartaigh, A. David Edwards, Joseph V. Hajnal, Mary Rutherford, Serena J. Counsell

## Abstract

**Background:** Altered structural brain development has been identified in fetuses with Congenital Heart Disease (CHD), suggesting that the neurodevelopmental impairment observed later in life might originate in utero. There are many interacting factors that may perturb neurodevelopment during the fetal period and manifest as structural brain alterations, such as altered cerebral substrate delivery and aberrant fetal hemodynamics.

**Methods:** We extracted structural covariance networks (SCNs) from the log Jacobian determinants of 429 in utero T2w MRI scans, (n = 67 controls, 362 CHD) acquired during the third trimester. We fit general linear models to test whether age, sex, expected cerebral substrate delivery and CHD diagnosis were significant predictors of structural covariance.

**Results:** We identified significant effects of age, sex, cerebral substrate delivery, and specific CHD diagnosis across a variety of SCNs, including primary motor and sensory cortices, cerebellar regions, frontal cortex, extra-axial CSF, thalamus, brainstem, and insula, consistent with widespread coordinated aberrant maturation of specific brain regions over the third trimester.

**Conclusions:** SCNs offer a sensitive, data-driven approach to explore whole-brain structural changes without anatomical priors. We used them to stratify a heterogenous CHD patient cohort, highlighting similarities and differences between diagnoses during fetal neurodevelopment. Although there was a clear effect of abnormal fetal hemodynamics on structural brain maturation, our results suggest that this alone does not explain all the variation in brain development between individuals with CHD.

## Introduction

Congenital heart disease (CHD) is the most common congenital malformation (EUROCAT 2020), occurring at a frequency of 0.8% to 1.2% of live births worldwide (Dolk et al., 2011, Bouma & Mulder 2017). CHD encompasses a wide spectrum of cardiovascular defects, from simple cardiac malformations to more complex and severe lesions that require surgical intervention. Following improvements in surgical and therapeutic intervention over the last two decades, most patients with CHD will survive into adulthood (Dellborg et al., 2023). However, many will display impairments affecting a wide range of neurodevelopmental domains (Latal et al, 2016, Newburger et al., 2012, Naef et al., 2017, Marelli et al., 2016, Bellinger et al 2011). Research into the mechanisms underpinning impaired neurodevelopment in CHD may enable appropriate preventative prenatal interventions to promote improved neurodevelopmental outcomes later in life.

With recent advances in fetal MRI, it has become possible to study brain development in utero. Previous work has identified deviations from normal brain growth trajectories in fetuses with CHD, including reduced regional brain volumes (Limperopoulos 2010, Clouchoux et al., 2013, Brossard-Racine 2014, Ren et al., 2021, Dovjak et al., 2022, Cromb et al. 2023), altered volumes of transient fetal compartments (Rollins et al., 2021, Wu et al., 2021), reduced cortical folding (Ortinau 2019, Jaimes et al., 2020) and a relationship between cerebral oxygen delivery and fetal brain size (Sun et al., 2015, Cromb et al, 2023). These studies demonstrate that abnormal neurodevelopment in the CHD population begins in utero during a period of rapid brain growth, featuring metabolically demanding cellular processes such as gyrification, oligodendrocyte maturation, and synaptogenesis. It has been hypothesised that aberrant cardiovascular physiology, altering the delivery of oxygen, glucose and other nutrients to the fetal brain during this critical window, impairs brain development (Peyvandi 2021, Lauridsen 2017, Sun 2015). Different types of CHD will affect the pattern of cerebral substrate delivery in different ways, likely having unique implications for brain development (Sun et al., 2021, Rollins et al., 2021, Cromb et al., 2023). There are also a multitude of other interacting factors that may perturb neurodevelopment during the fetal period and manifest as structural brain alterations. These include genetic factors, placental function, maternal stress, socioeconomic and environmental factors. Disentangling the effect of each of these factors remains a challenge, and therefore our understanding of structural brain abnormalities in the CHD population is incomplete.

In this work, we extracted structural covariance networks (SCNs) from the brain MRI of 429 fetuses. SCNs represent brain regions with coherent structural expansion and coordinated, independent maturational trajectories, often converging on functional brain networks (Geng et al., 2017, Seeley et al., 2009). SCNs have been used effectively to investigate changes in the organization of the brain at the network level, both in healthy ageing populations (Li et al., 2013, Geng et al., 2017, Spreng & Turner 2013), and in at-risk populations including preterm infants (Fenchel et al., 2021, Vanes et al., 2021), psychiatric patient cohorts (Bassett et al., 2008, Heinze et al., 2015), and in neurodegenerative disease (He et al., 2008, Seeley et al., 2009). SCNs offer a data-driven, dimensionality reduction approach, removing the dependency on predefined regions of interest, which allows the examination of dynamic contrast changes across space and time. This is particularly relevant for investigating the maturation of the fetal brain, which is rapid, complex, and characterised by the presence of transient structures (Kostovic & Rakic 1990). This model-free approach is also well suited to studying the heterogenous and varied pathophysiology of CHD, as it extracts features that represent independent modes of variation across the cohort. We hypothesized that by examining the sources of variance, we would be able to disentangle the effect of specific cardiac defects on the developing brain.

To address this challenge, we examined a large cohort of 429 fetuses, including 369 fetuses with CHD. There are many approaches to categorize CHD cohorts (Sun et al., 2015, Roberts et al., 2020) and explore the interplay between cardiovascular physiology and fetal brain development. For the purpose of this study, we divided the CHD cohort into groups based on the expected effect of the underlying cardiac defect on the streaming of substrate-rich placental blood to the cerebral circulation (Cromb et al., 2023). Each subject was reviewed individually, using fetal phase-contrast MRI flow data and/or contemporaneous echocardiography where available, as published in previous work (Cromb et al., 2023). This categorization was used to the test the specific hypothesis that cerebral substrate delivery affects the co-maturation of SCNs.

We also examined whether gestational age, sex, and CHD diagnosis are underlying sources of variation in structural brain maturation. With this approach, we stratified our large cohort of fetuses with CHD, identifying SCNs that vary across the population according to cerebral substrate delivery and more specifically, CHD diagnosis. We identify networks whose coordinated maturation is different from healthy controls but shared between multiple subtypes of CHD. We also find networks that are uniquely different for specific CHD diagnoses. Overall, this approach identified associations between specific cardiac defects and how they manifest in the developing fetal brain, which offers crucial insight into the potential structural underpinnings of neurodevelopmental abnormalities.

## Methods

### Ethical approval

The National Research Ethics Service West London committee provided ethical approval (07/H0707/105, 14/LO/1806, 17/LO/0292). Informed written consent was obtained before fetal MRI.

### Participants

Mothers were recruited between June 2014 and June 2022 at the Evelina Children’s Hospital in London, following routine ultrasound during the second trimester. 529 women (maternal age scan=31.52 (±5.73) years) were carrying a fetus with either known or suspected CHD. Participants were scanned during the third trimester, between 27 and 36 gestational weeks (GW). Exclusion criteria for mothers included multiple pregnancies, maternal weight over 125 kg (at time of scan), maternal diabetes, maternal hypertension, inability to give informed consent, or age under 18 years at the time of referral.

We also excluded fetuses with confirmed genetic diagnosis, such as 22q deletion syndrome, extracardiac anomalies, such as congenital diaphragmatic hernia or duodenal atresia, or structural brain abnormalities reported on fetal MRI, including bilateral ventriculomegaly, cerebellar hypoplasia or absence of the corpus callosum. After applying the exclusion criteria, the cohort consisted of 429 fetal MR scans (including 67 controls, and 362 fetuses diagnosed with CHD).

### Image acquisition & reconstruction

All scans were acquired on a Philips Ingenia 1.5T scanner, with 28-channel dStream anterior and posterior built-in coils. The T2-weighted fast-spin-echo sequence was specifically optimised for fetal imaging (TR = 13 ms, TE = 80 ms, image resolution= 1.25x1.25x2.5mm, slice thickness = 2.5 mm, slice spacing = 1.25 mm). A 3D slice-to-volume image reconstruction pipeline (available at https://github.com/SVRTK/SVRTK, https://hub.docker.com/r/fetalsvrtk/svrtk auto2.20) was used for motion correction, reconstructing the T2w images to 0.5mm3 isotropic resolution (Kuklisova-Murgasova 2012, Uus et al., 2021).

### MRI Quality control

All MRI scans were reviewed and reported by expert perinatal radiologists. The presence of any structural abnormalities was recorded, and image quality was scored on a scale from 1-4, based on the signal:noise ratio, presence of artefacts, field of view and residual motion (4 = High quality, 3 = Acceptable, 2 = Poor, 1 = Failed). Only subjects scoring 3 or 4 were included in the study, as previously described (Uus et al., 2022, Cromb et al., 2023).

### Cerebral substrate delivery categorisation

To explore the hypothesis that impaired cerebral substrate delivery plays a major role in the neurodevelopmental abnormalities associated with CHD, we divided the CHD cohort into 4 groups, according to the predicted level of substrate delivery to the developing fetal brain, based on the expected consequence of the underlying cardiac defect as described previously (Cromb et al, 2023).

A. Group 1 = Substrate content of cerebral blood is expected to be normal (n = 232 (113 male), μ = 31.81 GW ± 1.55),
B. Group 2 = Substrate content of cerebral blood is expected to be mildly reduced (i.e. some mixing of placental and fetal systemic venous blood) (n = 67 (35 male), μ = 32.37 GW ± 1.52),
C. Group 3 = Substrate content of cerebral blood is expected to be moderately reduced (i.e. complete mixing of placental and fetal systemic venous blood) (n = 46 (21 male), μ = 32.72 GW ± 1.74),
D. Group 4 = Substrate content of cerebral blood is expected to be severely reduced (i.e. complete reversal of normal placental streaming) (n = 23 (12 male), μ = 33.12 GW ± 2.02)

In the absence of direct measurements of cerebral substrate delivery, cases were classified by an expert fetal cardiologist, according to the expected effect of the underlying cardiac defect on the delivery of oxygen and nutritional content of blood to the carotid arteries, and by extension, the brain. MRI-derived fetal blood flows were used where available, as described in (Lloyd et., 2021). For cases where information about the direction of blood flow at the aortic isthmus could not be derived from MRI derived fetal blood flows, and was important for CHD categorization, it was extracted from the clinical fetal echocardiogram report, acquired as per routine clinical care. In cases where a diagnosis could potentially fit into more than one category, depending on severity or underlying hemodynamics, a combination of phase contrast (with metric optimized gating), fetal flow measurements (described in Lloyd et al., 2021, Janz et al., 2010) and/or contemporaneously acquired echocardiographic data were used to assign cases individually, following assessment of the data by a fetal cardiologist (Cromb et al., 2023).

### Image registration and Jacobian determinant calculation

Non-linear deformation fields were calculated to transform the native subject T2 to the age-matched template of the dHCP fetal atlas (https://doi.gin.g-node.org/10.12751/g-node.ysgsy1/) using ANTs Symmetric diffeomorphic image registration (Avants et al., 2008). Warps were then concatenated between native T2, the age-matched template and a 30 GW template space, which represents the median age of the cohort (Avants et al., 2008). The log Jacobian determinant was calculated on the concatenated warp, which represents the contraction and expansion of brain regions during image registration. In the resultant log Jacobian maps, higher log-Jacobian values represent brain regions that contracted during image registration (i.e., larger global and local brain volume), while smaller values represent smaller volume (Avants and Gee, 2004). The log Jacobian volumes for all subjects were concatenated to create a single 4D file, which was used as the input for the ICA (Van Rossum and Drake, 1995, Varoquaux et al., 2010).

### Independent Component Analysis (ICA) of Jacobian determinants

ICA is a data-driven, blind source separation technique that extracts salient patterns embedded in the data, it reduces the dimensionality of neuroimaging data (from many thousands of individual voxels) by separating out the multivariate signal into a maximally independent set of components (Jutten & Hérault, 1991). When applied in the spatial domain to structural imaging data (the log Jacobian determinants), ICA can detect coordinated growth between spatially separated brain regions, i.e. SCNs, which are strongly associated with other clinical or demographic variables (O’Muircheartaigh et al., 2014, Douad et al., 2014, Llera et al., 2019).

A canonical ICA algorithm (Varoquaux et al., 2010) was used, implemented in python using the nilearn package (Abraham et al., 2014). The ICA algorithm transforms the input data into components (or SCNs) that represent an ‘unmixing’ of the signal, such that the independent components have distributions that are non-Gaussian. The optimal number of SCNs (n = 40) for this case was chosen by surveying previous literature, and to balance robustness and interpretability (Eyre et al., 2021, Vanes et al., 2021). When the ICA dimensionality was increased above 40, visual inspection of components showed a division of cortical regions and splitting bilateral components into left/right lateralised areas.

The criteria for excluding components were (a) majority of the signal occurring in edge voxels, indicating misregistration (b) sparse, randomly distributed signal with low total area. For this dataset, all components passed the exclusion threshold and were included in subsequent analyses.

To extract weights, or ‘modes’, for each network in each individual subject, FSL’s general linear model was applied to the component maps and the Jacobian input volumes (Winkler et al., 2014, Anderson and Robinson 2001).

### Application of the General Linear Model (GLM) to identify covariates of brain structure

For each set of SCN weights, we fit a GLM, with SCN weights as the dependent variable, to test the hypothesis that age, sex, and cerebral substrate delivery were significant predictors affecting the modes of variation across the cohort.

We fit either 1A) or 1B) depending on whether the relationship between the SCN weights and GA was linear or 2^nd^ order polynomial (determined by the Akaike Information Criterion (AIC)).

> 1 A) Modes ∼ GA + sex + Cerebral substrate delivery
>
> B) Modes ∼ GA + GA^2^ + sex + Cerebral substrate delivery

We then used a subset of the cohort, to explore the effect of cardiovascular physiology more specifically for each CHD diagnosis. We selected diagnosis categories with > 10 subjects. These included Tetralogy of Fallot (ToF) (n = 13), Transposition of the Great Arteries (TGA) (n = 22), Right Aortic Arch (RAA) (n = 88), Hypoplastic Left Heart Syndrome (HLHS) (n = 25), Double Aortic Arch (DAA) (n = 27), Coarctation (CoA(+)) (n = 58), we also included fetuses with suspected coarctation prenatally who were not shown to have this condition in the neonatal period (“false positive” or CoA (-)) (n = 47). We included the CoA(-) group in this analysis, as it has been shown previously that this group differs significantly from a healthy control population in terms of the distribution of the fetal circulation, for reasons that remain unclear (Lloyd et al., 2021). Since the purpose of this work was to investigate the effect of cerebral substrate delivery on brain maturation, this group was included in our analysis.

> 2 A) Modes ∼ GA + sex + CHD Diagnosis
>
> B) Modes ∼ GA + GA^2^ + sex + CHD Diagnosis

We used FDR correction for multiple comparisons (between 40 models, one for each SCN) to adjust the p values for all predictors before reviewing whether they were significant in each model.

## Results

### Participants

429 subjects met the inclusion criteria, 67 were healthy controls (30 male) and 362 were diagnosed with CHD (182 male). All fetuses were scanned between 27- and 35-weeks gestational age (GA) (Figure 1). Histograms showing the distribution of GA at scan for each group are shown in Figure 1.

**Figure 1.**
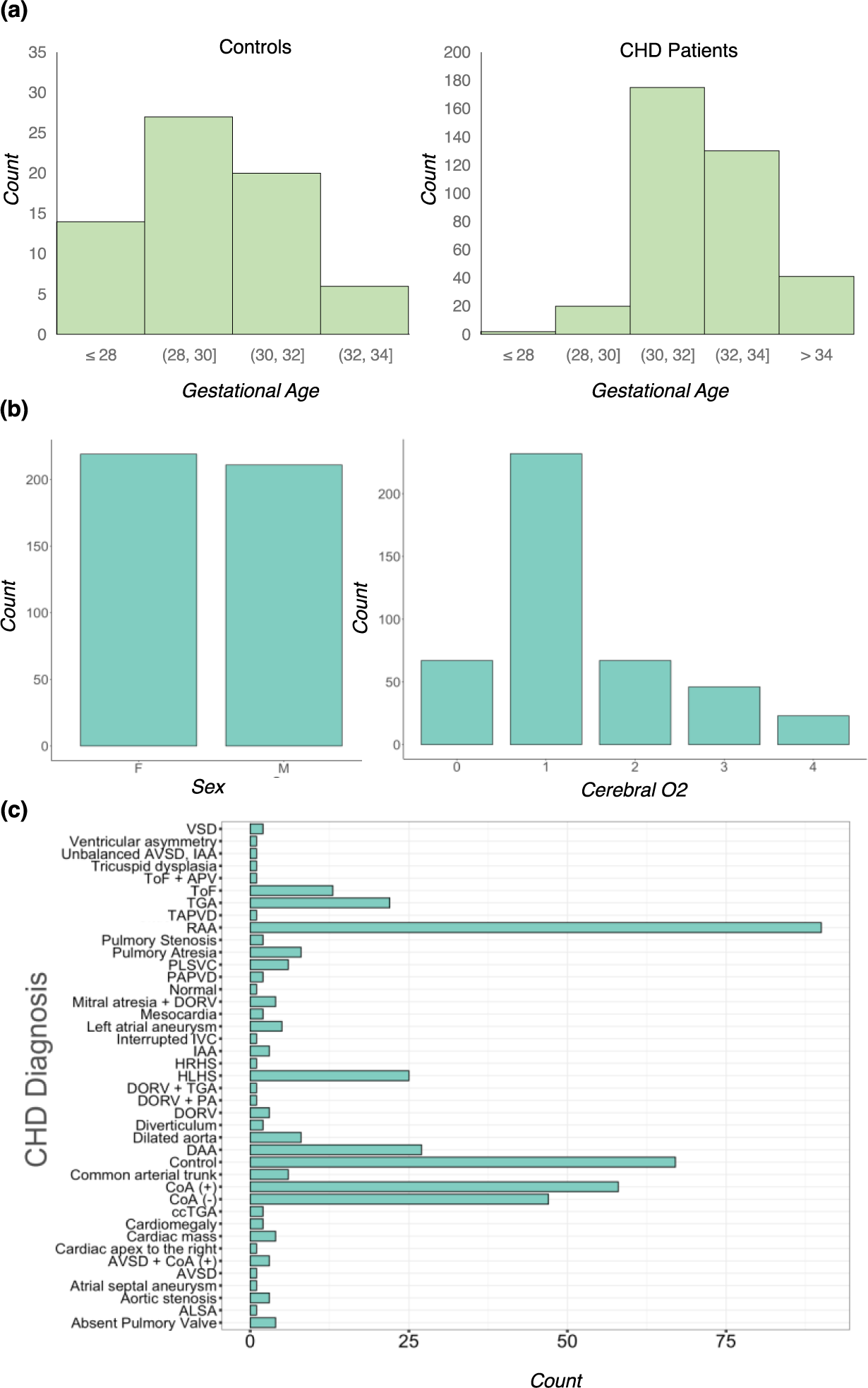
Distribution of covariates and cohort information. (a) Gestational age (GA) distribution for controls and CHD patient populations (b) Sex and Cerebral substrate delivery grouping (0 = control, 1 = expected normal, 2 = mildly reduced, 3 = moderately reduced, 4 = severely reduced). See Methods section ‘CHD categorisation’. (c) Distribution of CHD Diagnoses within the patient cohort.

A wide spectrum of CHD was represented in this cohort, including 40 different diagnoses (Figure 1, 2), which were categorised into 4 groups based on expected cerebral substrate delivery (Cromb et al., 2023), Normal = 232, Mildly reduced = 67, Moderately reduced = 46, Severely reduced = 23. The range of CHD diagnoses within each group is shown in Figure 2.

**Figure 2.**
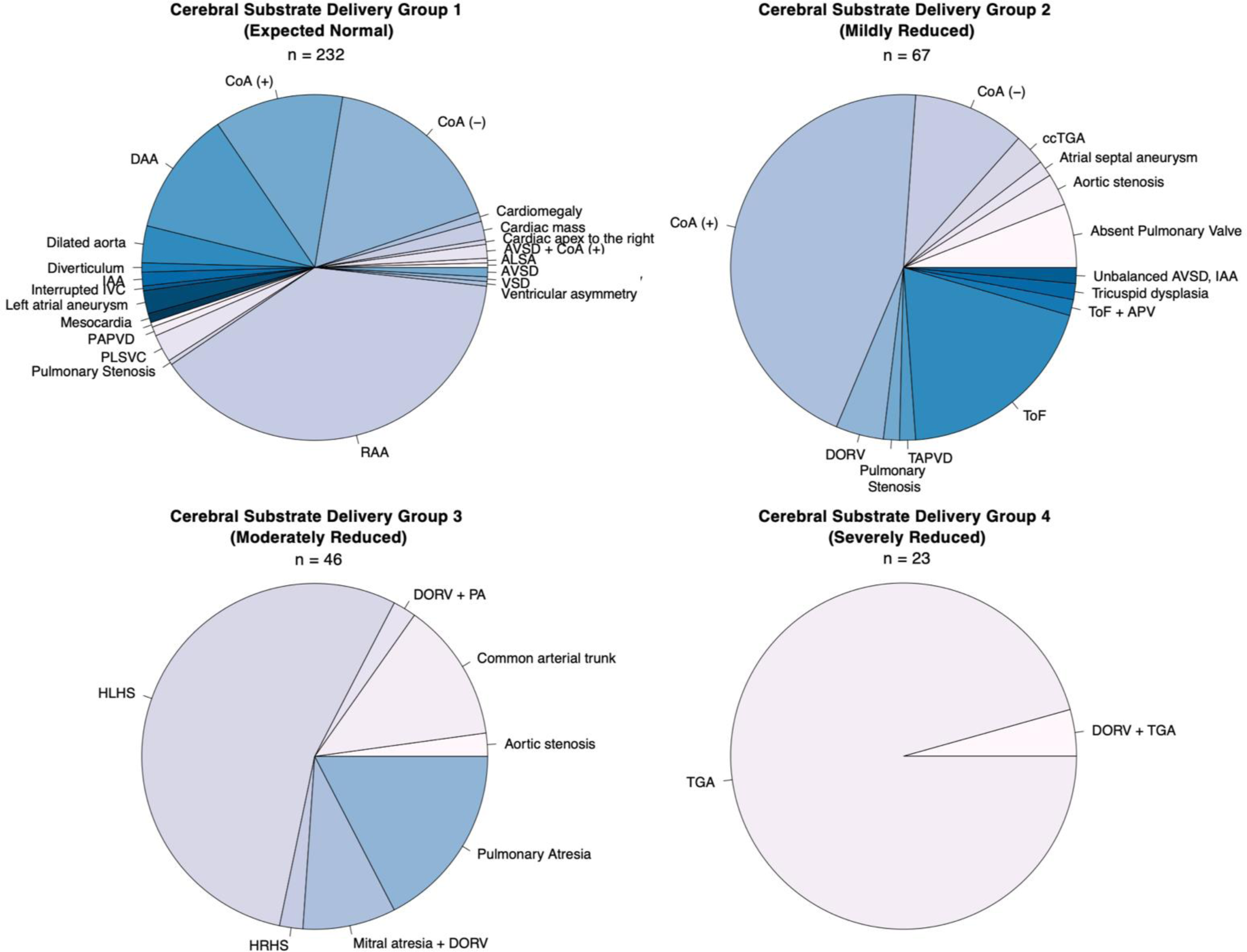
CHD grouping according to cerebral substrate delivery. ICA separates a multivariate signal (the T2w log jacobians) into additive, independent subcomponents that represent brain regions covarying between individuals.

### Structural covariance networks in the developing fetal CHD brain

As shown in Figure 3, we extracted 40 SCNs, each representing independent, coordinated structural development between brain regions (Figure 4). ICA was effective at extracting meaningful neuroanatomical structures, and labels were given to each SCN according to the corresponding anatomy (Figure 4). Almost all networks were either bilateral and symmetrical or had a complementary contralateral homolog component in the opposite hemisphere (Figure 4). All tissue types in the brain were represented, including deep grey matter, white matter, cortical grey matter, and CSF. Certain SCNs were isolated to a specific tissue, and others included multiple tissue types. Charting the SCN modes against gestational age shows general maturational trends (Supplementary info), however there is considerable variability between subjects of the same age, suggesting that there are other significant sources of variation between individuals, such as the abnormal fetal hemodynamics, for a large proportion of this cohort.

**Figure 3.**
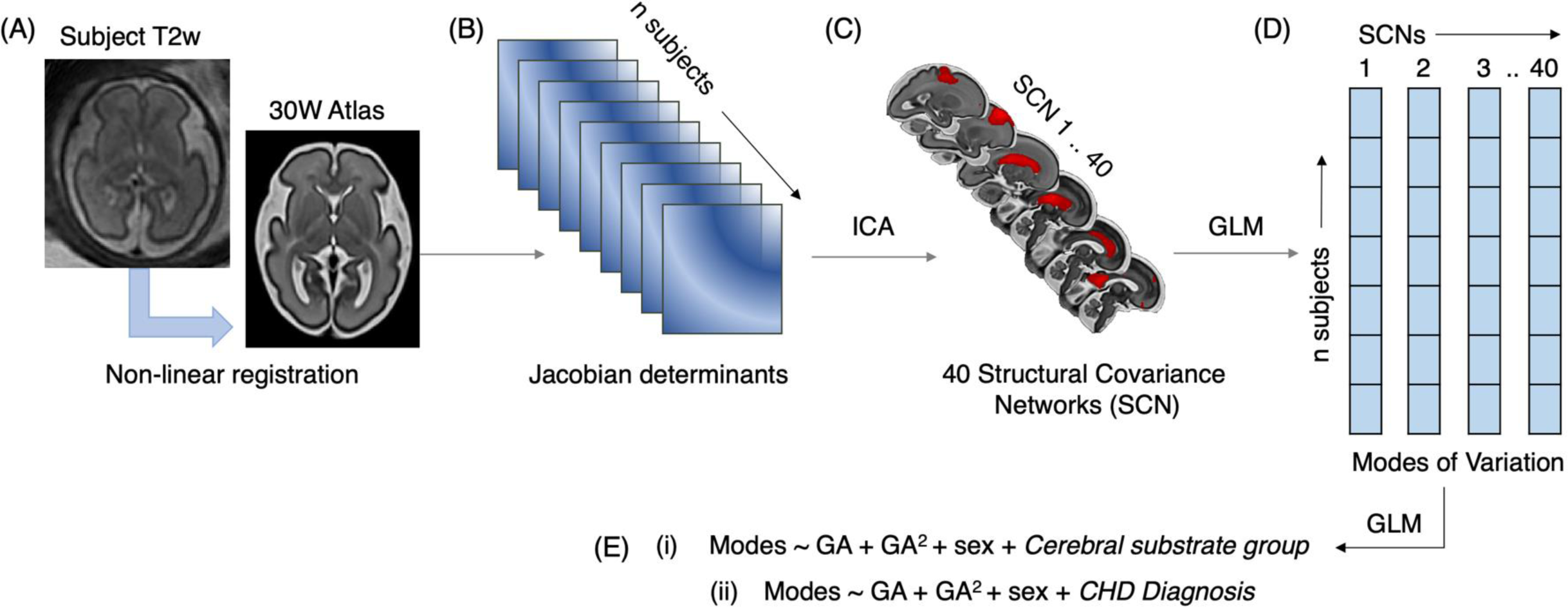
Methods pipeline to extract Structural Covariance Networks. (A) Reconstructed motion corrected T2w fetal MRI is registered to the 30 GW Atlas, using non-linear registration (B) The log Jacobian of the warp is calculated (C) Jacobians are concatenated across all subjects and used as input to the canonical ICA algorithm, to extract 40 SCNs (D) A GLM is fit to the Jacobians & the SCNs, to extract an SCN weighting (or ‘modes’) for each subject. (E) To examine the underlying sources of variation between modes for each SCN, two GLMs were fit, testing the effect of GA, GA^2^, sex and either (i) cerebral substrate grouping or (ii) CHD Diagnosis.

**Figure 4.**
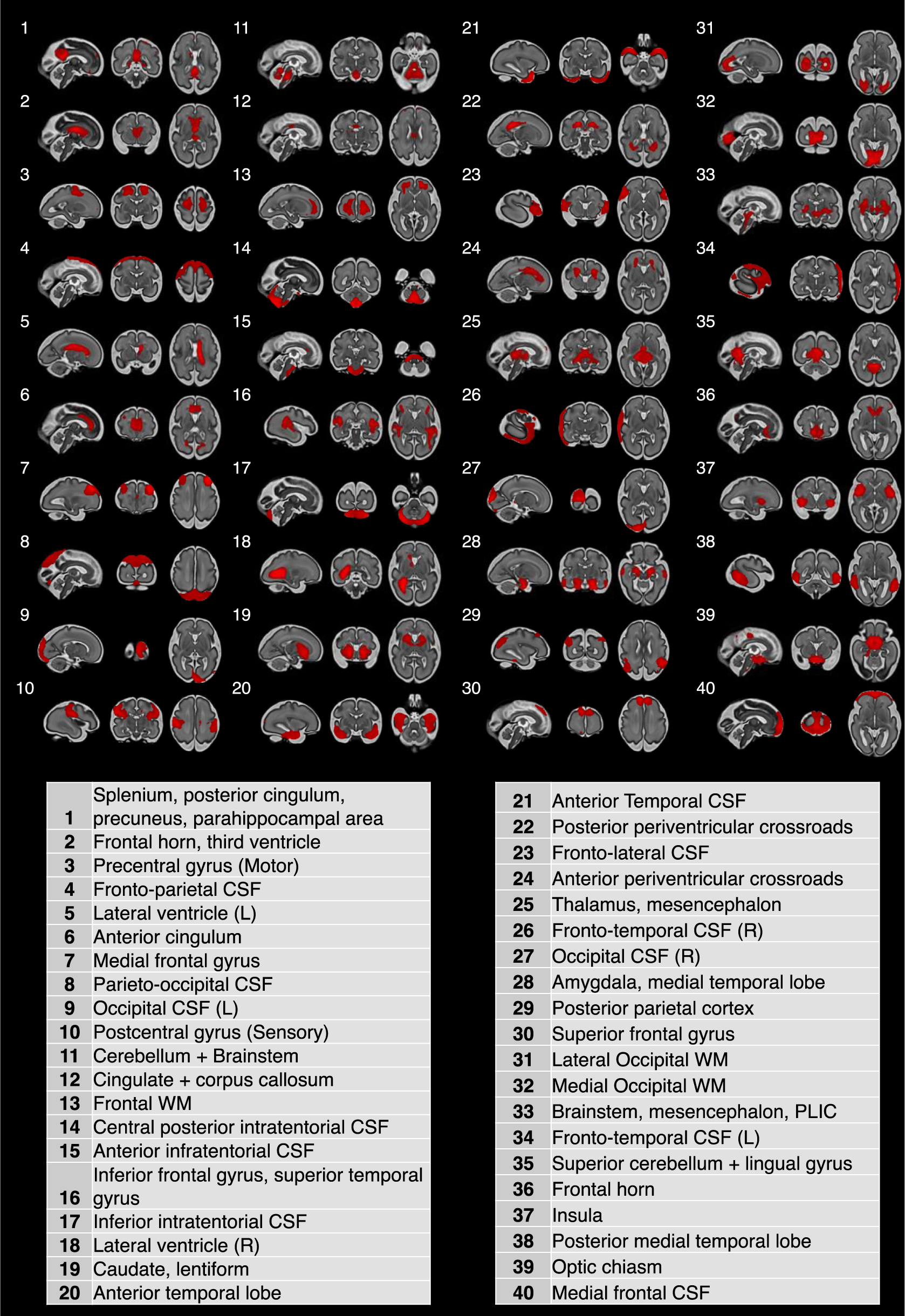
Structural covariance networks identified by ICA. 40 independent SCNs overlayed on a 30 GW fetal atlas. Table contains labels summarising their neuroanatomy.

### Structural covariance is associated with cerebral substrate delivery

We fitted a GLM to each set of component weightings (also referred to as ‘modes’) to test whether age, sex and cerebral substrate delivery were underlying sources of variation across the cohort. 35 SCNs were significantly associated with GA, 9 with sex and 14 with expected cerebral substrate delivery after FDR correction for multiple comparisons between 40 ICs (q < 0.05) (Table 1). When we examined each substrate delivery group compared to controls, we found significant variation between controls and CHD where cerebral substrate delivery was expected to be normal, in 3 SCNs, including the left and right frontal temporal cortex/CSF networks, and frontoparietal cortex. These networks were also significantly different between controls and groups 2 and 3 (mildly and moderately reduced). In addition, there were differences between controls and ‘mildly’ and ‘moderately’ reduced groups in the medial frontal gyrus. The ‘moderately reduced’ group were uniquely different from controls in the postcentral gyrus and optic chiasm, and different from the ‘expected normal’ group in these networks. The ‘moderately reduced’ group were also different from the ‘expected normal’ group in the anterior cingulum, thalamus, and mesencephalon networks.

**Table 1.**
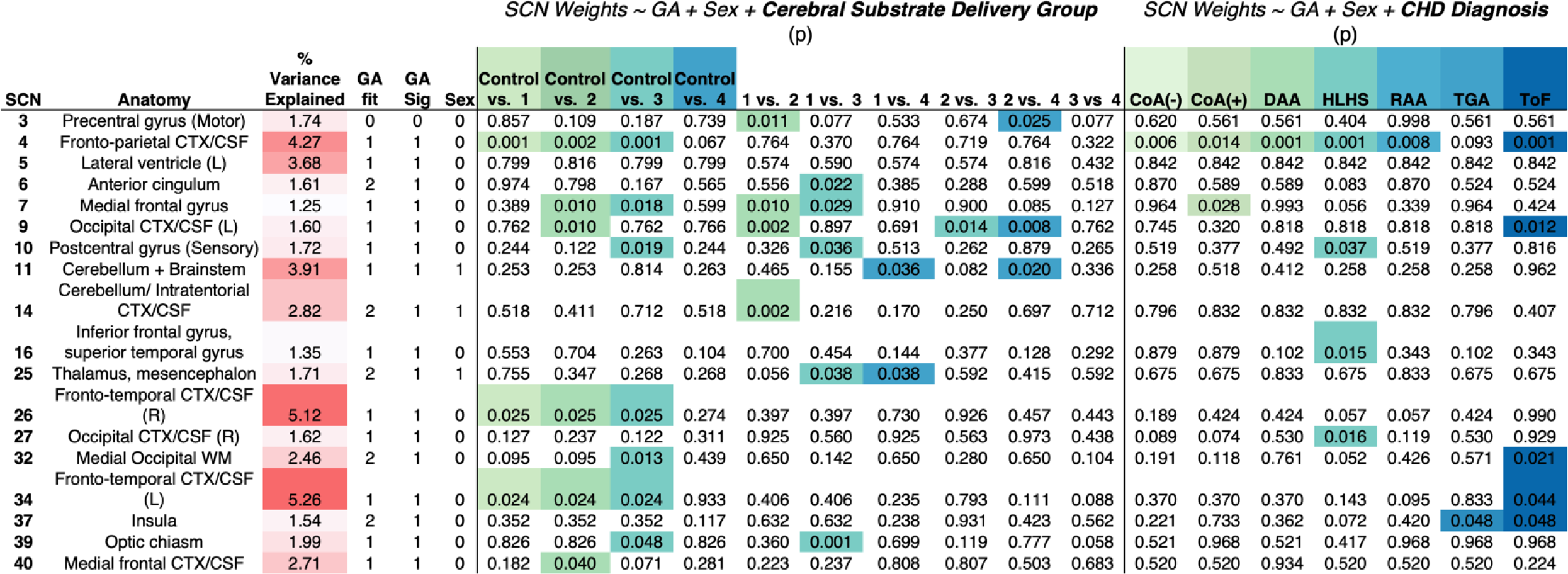
18 SCNs where Cerebral Substrate Delivery and CHD Diagnosis are significant predictors of variance. Adjusted p values after FDR correction for multiple comparisons between components (q < 0.05). Significant comparisons are indicated with coloured box. Table also contains column of the proportion of total variance across the cohort explained by each SCN, whether there was a relationship with GA, (0 = No relationship, 1 = linear, or 2 = 2^nd^ order polynomial), and if GA/sex were significant predictors of variance in the GLM.

There were no SCNs that have significantly different modes of variation between controls and the ‘severely reduced’ cerebral substrate delivery category. This might be due to a relatively small sample of fetuses in this category (n = 23), limiting our statistical power to detect differences in this group. However, we did find networks distinguishing the ‘severely reduced’ CHD group from the ‘expected normal’ and ‘mildly reduced’ groups. These networks included the precentral gyrus, occipital cortex/CSF, cerebellum & brainstem, and thalamus/mesencephalon. No significant differences were found between the ‘moderately’ and ‘severely’ reduced groups for any of the networks.

6 of the CHD-sensitive networks included high proportions of CSF, encompassing both extra-axial and infratentorial regions, suggested altered cortical expansion of these areas. These components also explained the highest proportion of the total variance across the cohort.

We created violin plots to show the model-fitted difference between groups, when accounting for age and sex, for components where there was a significant effect of expected cerebral substrate delivery (Figure 5). This highlighted the networks for which there was a gradient, or dose-dependent effect of cerebral substrate delivery, such as both left and right frontotemporal cortex/CSF networks and the cerebellum.

**Figure 5.**
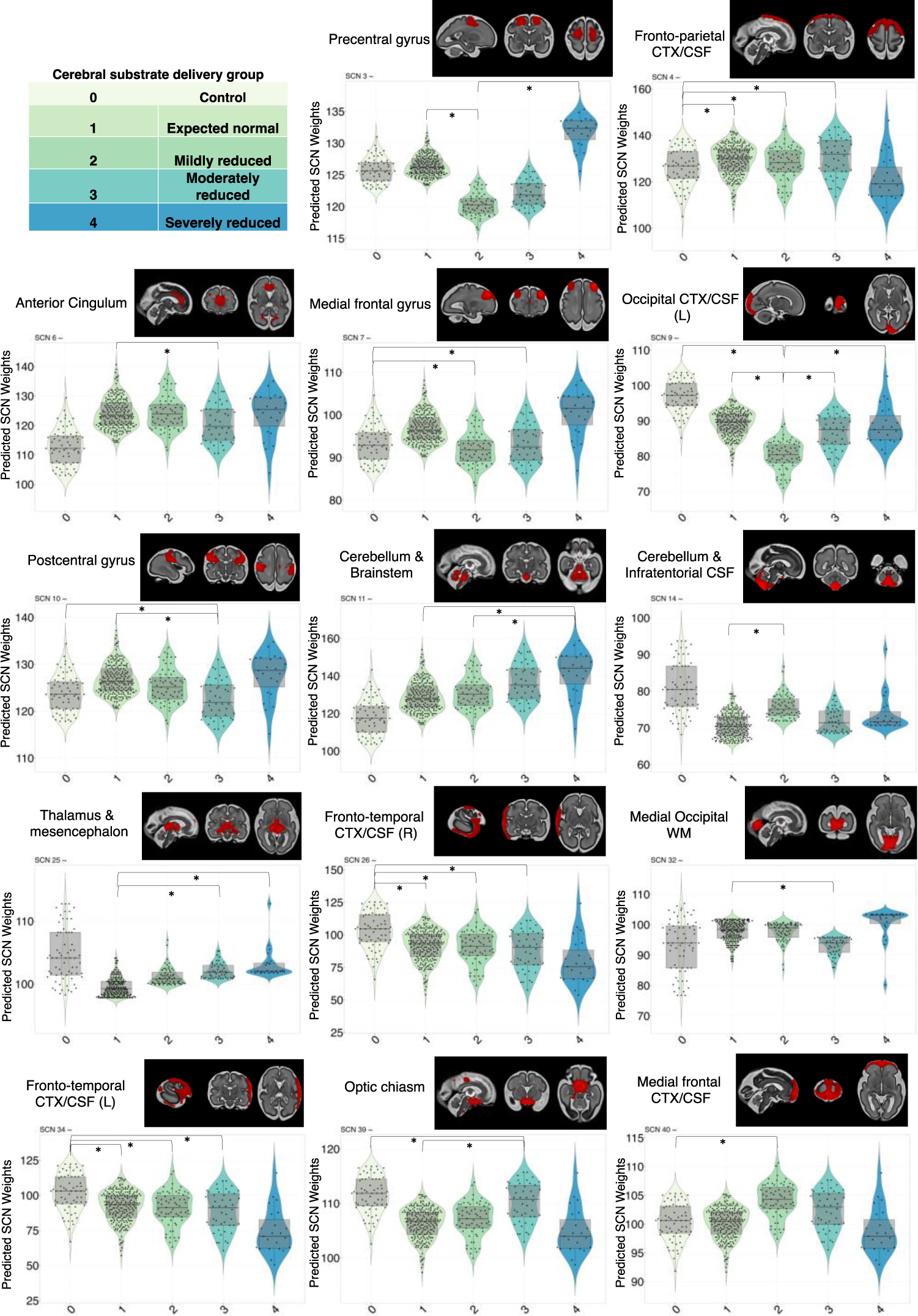
Structural covariance explained by cerebral substrate delivery. For each significant component, the GLM predicted modes are shown (when accounting for gestational age, sex and cerebral substrate delivery group), arranged along the x axis by cerebral substrate delivery groups (key shown in top left hand corner). (*) denotes significant difference between groups after correcting for multiple comparisons (FDR, q < 0.05).

### The effect of specific CHD diagnoses on structural brain maturation

Given the differences in structural covariance across the brain between CHD cerebral substrate groups, we carried out a sensitivity analysis in a subset of the cohort, to explore whether specific CHD diagnoses were predictors of variation (Table 1). In this way, we could test the hypothesis that specific cardiac defects have unique and distinct effects on structural brain maturation. We used a subset of the cohort (n = 238), only including CHD diagnosis categories with > 10 subjects (see Methods). We fit a GLM with SCN weights as the dependent variable, and tested for a combined effect of age, age^2^, sex and CHD diagnosis. There were 9 different SCNs where at least one of the CHD diagnosis categories explained a significant amount of the variation between individuals. The summary of SCNs that were significant for each diagnosis category after FDR correction (q < 0.05) are shown in Figure 5.

We found the frontoparietal network was significantly different between controls and all CHD diagnoses except for TGA (Table 1). We also found certain SCNs, where only one CHD diagnosis group was significantly different from controls. CoA (+) explained a significant proportion of the variation in the network corresponding to the medial frontal gyrus. HLHS explained a significant proportion of the variance in the postcentral gyrus, inferior frontal & superior temporal gyrus, and occipital cortex/CSF. Fetuses with ToF were uniquely different from controls in the frontotemporal cortex, medial occipital white matter, and occipital cortex. Both ToF and TGA were significant predictors of variance in the insula network.

## Discussion

In this study we applied an unsupervised, data-driven analysis technique, ICA, to capture the dynamic T2w contrast changes across the fetal brain in space and time, summarising them into an interpretable set of anatomically meaningful networks, or SCNs (Comon 1994). The networks we derived correspond to recognisable structures in the fetal brain, including regions that are developmentally critical but transiently present in the fetal period, such as anterior and posterior periventricular crossroads. We used the networks to stratify the large heterogenous cohort of different CHD cases, identifying variation across the brain according to cerebral substrate delivery and specific CHD diagnosis. Of the 40 SCNs we derived, we found a significant effect of cerebral substrate delivery or CHD diagnosis in 18 of them.

### The maturation of specific networks is vulnerable to the effects of reduced cerebral substrate delivery

Previous work has demonstrated that hemodynamic alterations in fetuses with CHD contribute to abnormalities in brain development (Limperopoulos et al., 2010, Sun et al., 2015). Volumetric differences correlate with cerebral substrate delivery in the third trimester (Peyvandi et al., 2021, Cromb et al., 2023), which represents a critical window of accelerated fetal brain growth and increased cerebral metabolic demands. A reasonable hypothesis to explain these findings is that reduced cerebral substrate delivery, secondary to altered fetal cardiovascular physiology, is a driving factor in the emergence of structural brain differences in CHD (Rudolph, 2010). In this study, we identified networks of brain regions vulnerable to the effects of altered cerebral substrate delivery which can be broadly grouped as follows, (1) Gyral areas, pre- and post-central, medial frontal (2) Thalamus & brainstem (3) Cerebellum & infratentorial CSF, (4) Cortical/CSF regions, frontoparietal, frontotemporal and occipital cortex/CSF. The sensitivity of these particular regions is congruent with results of other studies, also noting the most significant volumetric changes in CHD in the frontal lobe and the brainstem (Ortinau et al., 2019), impaired expansion of higher cortical areas (Leonetti et al., 2019), altered patterning and delayed maturation of cortical folds (Ortinau et al., 2019, Dovjak et al., 2022, Clouchoux et al., 2013, Kelly et al., 2017), abnormal cerebellar development (Dovjak et al., 2020) and enlarged CSF spaces (Limperopoulos et al., 2010, Jorgensen et al., 2018, Schellen et al., 2015, Mlczoch E et al., 2013, Brossard-Racine et al., 2014, Ng et al, 2020). Studies across multiple different cohorts, spanning a large gestational age range, between 20 weeks GA and term, have reported increased extra-axial CSF spaces in the fetal brain with CHD (Limperopoulos et al., 2010, Jorgensen et al., 2018, Schellen et al., 2015, Mlczoch E et al., 2013, Brossard-Racine et al., 2014), which has been interpreted as a general marker of delayed brain development in this population (Brossard-Racine et al., 2014).

### Cerebral substrate delivery explains some but not all of the variance in CHD

We observed differences between healthy controls and CHD fetuses where cerebral substrate delivery is expected to be normal, in frontoparietal, frontotemporal and occipital cortical/CSF networks, suggesting extrinsic factors to this analysis, such as genetic and environmental factors, also mediate the differences in early brain growth. Furthermore, when we explored specific CHD diagnoses in a subset of the cohort, we recapitulated this result, finding CoA (-), RAA, and DAA subtypes predicted the variance in the same frontoparietal cortex/CSF networks. These diagnoses are considered milder forms of CHD where cerebral substrate delivery is expected to be normal (Rudolph 2010). An alternative explanation for the difference in brain development is genetic variation among the milder phenotypes, which has not been accounted for in this analysis. Previous work has highlighted shared underlying genetic pathways between the heart and brain which may account for the phenotype of altered structural brain development in CHD (Unolt et al., 2018, Richards & Garg 2010). Genetic abnormalities are also highly prevalent in the CHD population (Blue et al., 2017), and protein-damaging *de novo* mutations have been identified for genes highly expressed in both the developing heart and brain (Homsy et al., 2015, Ji et al., 2020).

The residual variation in brain development between individuals in this work may also be mediated by differences in placental function. The parallel development between fetal and maternal organs, and the shared genetic and developmental pathways between the heart, brain, and placenta, are emerging as important contributing factors to the vulnerability of this patient group (Steinweg et al., 2021, Rychik et al., 2018, Jones et al., 2015, Matthiesen et al., 2016). Placental imaging studies have highlighted a critical relationship between placental size and overall fetal growth, in both healthy and at-risk populations, suggesting that over a third of birth weight variation is due to placental weight (Salafia et al., 2008). A growing body of evidence implicates abnormal placental structure and function in CHD pathology (Steinweg et al., 2021, Rychik et al., 2018, Jones et al., 2015, Matthiesen et al., 2016), and future work would benefit from investigating these effects on the structural brain.

### Specific diagnoses were better predictors of variance than others

When examining the differences between CHD diagnoses, HLHS and ToF were the most significant predictors of variance across different networks, in 4 and 5 SCNs respectively. We found a unique effect of HLHS diagnosis on three structural networks, including the postcentral gyrus, inferior frontal and superior temporal gyrus, and occipital cortical/CSF. In ToF fetuses, the medial occipital white matter, the insula, and the frontotemporal and occipital cortex were also significantly different from healthy controls. Previous studies have also observed a severe effect on brain development in fetuses and neonates diagnosed with either ToF (Schellen et al., 2015) or HLHS (Glauser et al., 1990, Clouchoux et al., 2013). One study reported significantly reduced cardiac output in HLHS fetuses and a dose-dependent effect on delayed brain maturation (Sun et al., 2015). Our analysis also highlighted the insula network as significantly different for fetuses with ToF or TGA compared to controls, in accordance with previous work noting delayed opercular development in term infants with complex CHD (Glauser et al., 1990, Masoller et al., 2016, Ortinau et al., 2019, Peng et al., 2016). Operculation of the insula is usually complete at term (Goldstein et al., 2017), however an open operculum and exposed insular cortex have been associated with neurodevelopmental delays in CHD and more broadly in other patient populations (Mahle et al., 2000, Licht et al., 2009, Tatum et al., 1989, Chen et al., 1996). Interestingly, TGA was not significantly different from controls for any of the networks except the insula. Although previous work has shown that whole brain volume is reduced in fetuses with TGA, and disproportionately smaller than the volume of the fetal body (Jorgensen et al., 2018, Cromb et al., 2023), our results suggest that the local effects across the brain are minimal. It is plausible that although the global brain size is smaller for this subtype, coordinated growth at the local level between brain structures is normal.

### CHD-sensitive networks in frontal cortical regions

Specific networks emerged as being affected by both cerebral substrate delivery and CHD diagnosis categories. These included frontoparietal, frontotemporal and occipital cortex. In the fetal period, these networks likely represent altered volumes of CSF, in conjunction with the areal expansion and gyrification of the cortices. Previous work using a porcine model, explored the developmental processes effected by both transient and chronic disturbances in fetal oxygen delivery (Xuegang et al., 2011, Ishibashi et al., 2012, Morton et al., 2017), resolving that this may provide a mechanistic explanation for the effect of CHD on the development of higher order cortical areas. They established that inducing chronic hypoxic exposure decreases neuronal proliferation, migration, interneuron populations and overall volume in the insula and prefrontal cortices (Morton et al., 2017). In the same study, a parallel analysis of post-mortem brain tissue from infants with complex CHD revealed less mature astrocytic processes, and a depletion of neuroblasts within the subventricular zone in frontal areas (Morton et al., 2017). The authors speculate that this cellular phenotype may propagate to the level of neuronal circuits, creating an excitatory/inhibitory imbalance in the developing cortex in CHD (Morton et al., 2017). Excitatory/inhibitory imbalance has been investigated across a wide spectrum of intellectual and behavioural disabilities, potentially explaining why children with CHD show deficits in cognitive domains associated with higher order cortices (Leonetti et al., 2019).

### White matter dominated networks were largely not affected by CHD diagnosis

Many white matter networks were extracted by this analysis, reflecting the growth of key white matter structures and development of fibre connectivity over the third trimester (Kostovic & Milosevic 2006, Wilson et al., 2021), including both the anterior and posterior periventricular crossroads, and the corpus callosum/cingulum components. However, in most of these networks we did not detect significant differences between CHD and the control population. Previous studies characterising white matter development in utero in fetuses with CHD (Clouchoux et al., 2013, Ortinau et al., 2018), show smaller white matter volumes, which may just reflect the overall reduction in brain size in this group. Diffusion weighted imaging (DWI) may be more appropriate to elucidate any aberrant white matter development in CHD, as changes can be examined at the microstructural level. To the best of our knowledge, there is one previous study using in utero DWI in CHD, which suggests a group level reduction in fractional anisotropy in the body and splenium of the corpus callosum (Khan et al., 2018). However, a larger cohort of patients over a wider gestational age range is necessary to improve understanding of group level differences, as diffusion metrics show a non-linear relationship with gestational age in fetal white matter (Wilson et al., 2021).

## Conclusions

Overall, this analysis framework offers an alternative approach to studying a heterogenous patient group, capturing variation associated with age, cerebral substrate delivery and CHD diagnosis. This work contributes to building a more comprehensive understanding of brain development in CHD before birth, in utero, highlighting regions that maturing differently in this vulnerable population. The significant effect of cerebral substrate delivery and CHD diagnosis on deep grey matter, cortical regions, cerebellum and CSF supports the view that a lower oxygen environment can lead to adverse neurodevelopment. However, we also demonstrate that there is a lot of variation between individuals not attributable to fetal hemodynamic alterations, supporting the hypothesis that in utero neurodevelopment in CHD is impacted by a complex combination of factors, including genetics, maternal stress, and placental function. Our unconstrained data-driven approach identified the same vulnerable brain regions as previous work (Ortinau et al., 2019, Dovak et al., 2022, Clouchoux et al., 2013, Limperopoulos et al., 2010, Jorgensen et al., 2018, Schellen et al., 2015, Claessens et al., 2019, Mlczoch E et al., 2013, Brossard-Racine et al., 2014, Glauser et al., 1990, Masoller et al., 2016, Peng et al., 2016), which was reliant on a-priori segmentations. The reproducibility of this result supports that these findings are a meaningful reflection of biological differences between the patient and control group. The results highlight the potential for neuroimaging data in the fetal period to provide biomarkers for CHD subtypes, disentangling the unique effects of different CHD diagnoses on structural neurodevelopment. Earlier identification of the structural brain networks that confer neurodevelopmental risk increases the likelihood of successful targeted intervention to improve outcomes later in life.

## Limitations

There are some important limitations to note with this study. Firstly, the distribution of gestational ages between controls and fetuses with CHD was different. Although our model included GA, this may still result in some false positive or false negative significant networks. We were unable to discern the delivery of specific substrates, or measure substrate delivery quantitatively for every fetus. The cerebral substrate delivery groups also had uneven sample sizes, with a much higher proportion of subjects in the ‘expected normal’ group than any other group. This gave us more statistical power to detect differences between group 1 and controls compared to other groups, which may have led to some false negatives, as we would expect to find more differences between controls and more severe forms of CHD (Group 4 cases). Similarly for CHD diagnoses, more severe subtypes (TGA, ToF and HLHS) were less common, therefore our statistical tests were under powered, hampering our ability to detect differences. All imaging data was acquired on the same scanner, in the same hospital catchment area in central London, and despite the socioeconomic and racial diversity of London, our result may not be generalizable to other populations.

## Data Availability

Restrictions apply to the availability of these data, which were used under license for this study. Certain anonymised data are available upon request from the corresponding author.

## Acknowledgments

We would like to thank all the families who participated in this research. We would also like to thank the research radiologists, radiographers, and staff from the Evelina London Children’s Hospital Fetal and Paediatric Cardiology Departments and the Centre for the Developing Brain at King’s College London. Ethical approval for this study was provided by The National Research Ethics Service West London (07/H0707/105, 14/LO/1806, 17/LO/0292).

